# Using discrete- and continuous-time machine learning models (Nnet, CoxNet, GLMnet) to explore sex and age differences in stroke prediction among hypertensive individuals

**DOI:** 10.1101/2025.08.22.25334217

**Authors:** Gideon MacCarthy, Raha Pazoki

## Abstract

**Introduction:** Stroke is one of the leading causes of death and long-term disability globally. Several studies have investigated the incidence and predictors of stroke in the healthy population; only a few have specifically focused on stroke risk prediction among individuals with hypertension. Given that hypertension is the most common modifiable risk factor for stroke, this represents an important research area to study. Improving our understanding of stroke risk among hypertensive individuals has the potential to significantly improve preventative efforts and decrease the worldwide stroke burden.

**Material and Methods:** The current study involved 116,216 participants of European ancestry from the UK Biobank. We created stroke genetic liability using data from two predictive models using clinical, lifestyle, and genetic factors (stroke genetic liability) in the training set, namely a Cox proportional hazard (CoxPH) model, a penalized Cox proportional hazard model (CoxNet), a penalized logistic regression model (GLMnet), and a neural network (Nnet), to estimate time-to-event risk for stroke among hypertensive patients, older and younger hypertensive patients, and hypertensive males and females separately. We then assessed their performances in the testing set with the area under the curve (AUC) and Brier score (BS).

**Results:** The CoxPH, CoxNet, and GLMnet achieved an equal AUC of 67.0% in hypertensive patients. All models demonstrated superior performance in men compared to women, with the CoxPH model achieving the best AUC of 68% in men. All models demonstrated superior performance in older patients compared to younger patients, with the GLMnet achieving the best AUC of 61% in this group.

**Conclusion:** Including stroke genetic liability in prediction models slightly improves stroke prediction for hypertensive men and older patients. The Cox proportional hazards (CoxPH) model outperformed the machine learning models in predicting stroke risk among hypertensive men.

## INTRODUCTION

Stroke is ranked among the leading causes of mortality and long-term disability in adults worldwide [1, 2]. Over 1.2 million stroke survivors live in the United Kingdom (UK). Every year, more than 100,000 people living in the UK suffer from a stroke. Between 2015 and 2035, stroke incidence is projected to increase by 60% and stroke prevalence by 120% annually [3].

Studies have shown that several risk factors, both modifiable, such as hypertension, diabetes mellitus, and chronic kidney disease, and non-modifiable, such as genetic factors, age, and sex, play a significant role in the complex mechanism of stroke occurrences [4].

According to evidence from many studies, hypertension is the most common risk factor for all types of strokes, and the majority of stroke patients have a history of hypertension [5].

Hypertension significantly increases the risk of stroke in two main ways. It damages the walls of small blood vessels in the brain, causing them to become narrower, stiffer, and more prone to plaque buildup or rupture. These damaged vessels can either lead to ischemic stroke, caused by plaque blocking blood flow to the brain, or hemorrhagic stroke, caused by the rupture of a weakened blood vessel (www.stroke.org.uk/stroke/managing-risk/high-blood-pressure, accessed on 13/08/25).

Stroke risk increases with age and varies by sex and the presence of other risk factors, such as hypertension, diabetes mellitus, atrial fibrillation, lipids, cigarette smoking, physical inactivity, chronic kidney disease, and family history [6]. Men are more likely than women to experience a stroke at a younger age. However, as women get older, especially beyond 75 years, their risk of stroke becomes higher than that of men [5, 7].

Several stroke prediction tools, including the two widely used models, the Framingham Stroke Risk Score (FSRS) and the Ischemic Cardiovascular Disease model (ICVD), have been developed with non-genetic risk factors such as age, sex, smoking, blood pressure (BP), total cholesterol, diabetes mellitus, heart disease, and body mass index (BMI) to predict a person’s 10-year risk of stroke [8]. However, the two models do not integrate genetic liability into the risk estimate or include complex, nonlinear interactions among risk factors.

Recently, machine learning models have been increasingly applied to predict the risk of stroke, providing the ability to capture complex, nonlinear relationships among risk factors that traditional statistical models might not detect. Several studies [9–13] have developed machine learning models for risk of stroke prediction. These models, including the random forests, neural networks, decision trees, and gradient boosting machines, have been compared to traditional statistical approaches such as the Cox proportional hazards model. However, there is no consistency on which prediction model is a better fit. While some studies have demonstrated superior prediction performance of machine learning models over traditional models in certain datasets, others have found only a marginal or no advantage, or the reverse. This inconsistency may be attributed to the difference in study design, study population, etc. As a result, no single machine learning or traditional model has appeared as consistently superior across studies.

Although several studies have investigated the risk and predictors of stroke, only a few have specifically focused on stroke risk prediction among individuals with hypertension [14–16]. There is limited or no evidence on stroke risk prediction models for hypertensive European populations, specifically using large-scale cohorts such as the UK Biobank. Considering that hypertension is the most common modifiable risk factor for stroke, this limitation represents an important research area to investigate. Furthermore, this research gap suggests the need for population-specific models that can improve personalized stroke risk assessment and prevention strategies in hypertensive individuals. Therefore, in this study, we aim to develop and evaluate a stroke risk prediction model for hypertensive European populations. The current study will contribute to improving the prediction of stroke risk, specifically among individuals with hypertension. Our findings may contribute to the efforts of making more accurate stroke risk classification, thereby informing clinical decision-making and public health policies driven by the reduction or prevention of hypertension-related stroke

The current study has four main objectives, including (1) assessing the predictive value of stroke genetic liability in the prediction of stroke in hypertensive individuals, (2) assessing the predictive value of stroke genetic liability in the prediction of stroke among hypertensive men and women separately, (3) assessing the predictive value of stroke genetic liability in the prediction of stroke among older and younger hypertensive patients, and (4) comparing the performance of the Cox proportional hazard model and machine learning models within the full continuous follow-up time versus discrete follow-up time.

## METHODS AND MATERIALS

### Ethical Approval

The UK Biobank (UKB) received ethical approval from the Northwest Multi-Centre Research Ethics Committee as a Research Tissue Bank, and all participants provided written informed consent at the time of recruitment. The current study was conducted using UK Biobank data under approved application number 60549. Additionally, ethical approval to use secondary data from the UK Biobank for the current study was obtained from the College of Health, Medicine, and Life Sciences Research Ethics Committee at Brunel University of London (reference: 27684-LR-Jan/2021-29901-1).

### 2.2 Source of Study Population

Participants in this study were of European ancestry and recruited from the UK Biobank, a large, prospective cohort study. The design, recruitment procedures, and data collection methodologies have been described in detail in our previous publications [12, 17] and by Sudlow [18]. In brief, the UK Biobank recruited over 500,000 individuals aged 40–69 years between 2006 and 2010 from 22 centres across the UK. Baseline data included sociodemographic factors, lifestyle, medical history, physical measurements, and linked health records from Hospital Episode Statistics (HES) and national registries. The current study is based on hypertensive participants in the UK Biobank.

### Inclusion Criteria

The current study included participants who had complete data on all relevant confounding variables. Individuals were classified as having hypertension if they met any of the following criteria: (i) a self-reported physician diagnosis of hypertension, (ii) a measured systolic blood pressure (SBP) ≥ 140 mmHg or diastolic blood pressure (DBP) ≥ 90 mmHg at baseline, or (iii) a record of using blood pressure-lowering medication either before or at the time of recruitment

### Exclusion Criteria

Participants were excluded from the study based on the following criteria (**Figure 1**):

- Pregnant women and those uncertain of their pregnancy status (N = 278).
- Individuals with mismatches between self-reported and genetically determined sex (N = 320).
- Participants related up to the second degree were identified using a kinship coefficient cutoff of 0.0884 (N = 33,369).
- Individuals with a prior or current diagnosis of vascular or heart problems reported at baseline (N = 25,340).
- Participants using cholesterol-lowering medications (N = 34,243).
- Individuals who reported ceasing smoking or alcohol consumption due to health reasons or medical advice (N = 58,752).
- Participants with missing data on key confounding variables (N = 61,961).
- Non-hypertensive participants were defined as those not meeting the hypertension criteria outlined in the inclusion section.

**Figure 1.**
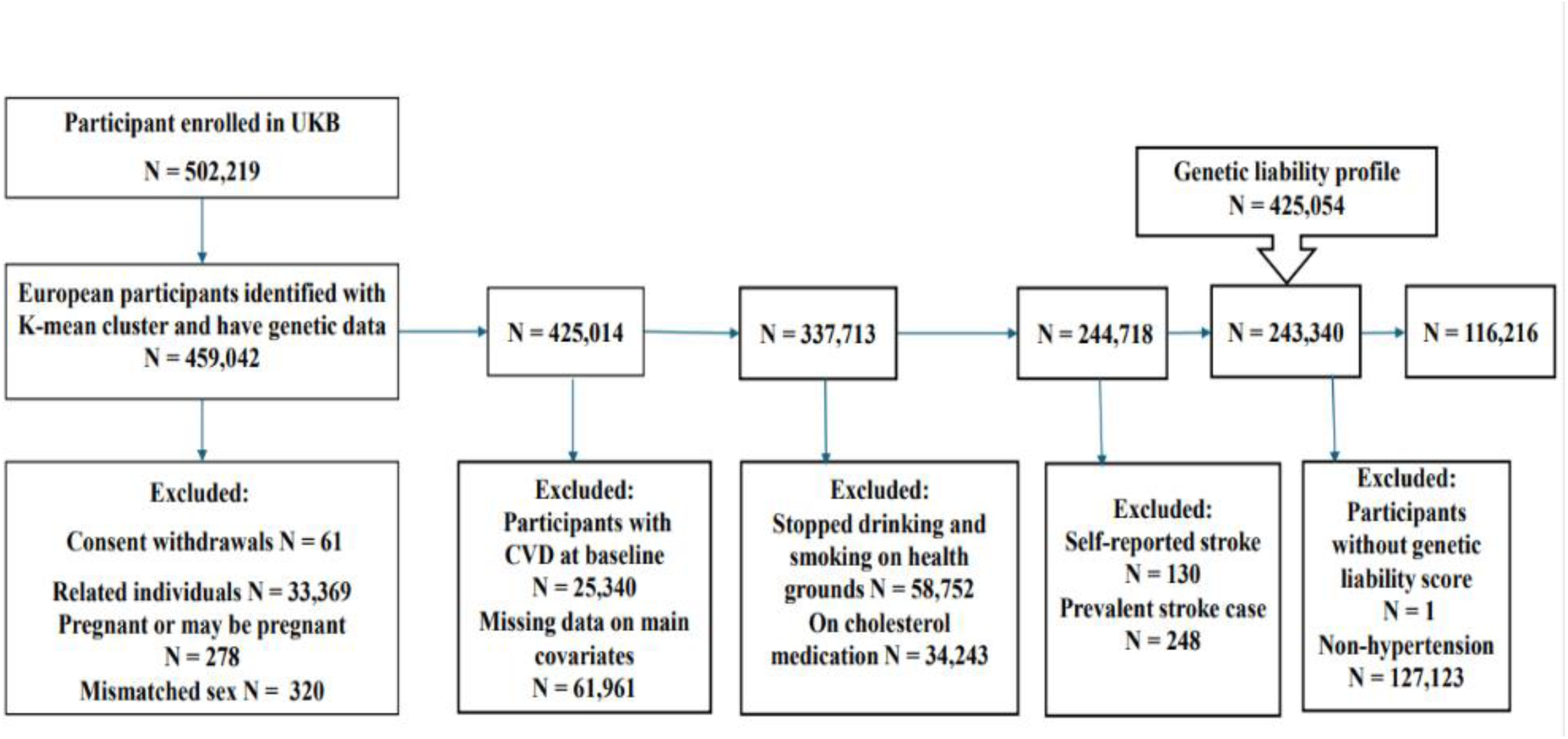
Exclusion Criteria of the study: The flowchart of the study participant selection. UK Biobank (UKB) data had over 500,000 participants at the time of the beginning of this study. We used the K-mean cluster method to extract 425,054 participants of European ancestry and have genetic data. The final dataset included 116,216 participants who met the inclusion criteria.

After applying these criteria, a final analytic sample was retained for downstream analysis.

### Study Population and Period

This study focused on a subset of unrelated UK Biobank participants of European ancestry (N = 116,216; **Figure 1**) who met criteria for hypertension at baseline. The follow-up period for this study extended from the date of each participant’s baseline health assessment (conducted between 2006 and 2010) to the end of March 2017. Participants who did not experience a stroke event by the end of the follow-up period were censored at that time.

### Genotyping and Imputation

Details of DNA extraction, genotyping, and imputation procedures have been described in our previous manuscript [12, 17] and [19–21]. Briefly, genotyping was performed by the UK Biobank using the UK Biobank Axiom array. Genotype imputation was conducted using the IMPUTE4 software with reference to the UK Biobank’s centrally estimated genetic principal components and kinship coefficients, in accordance with the Haplotype Reference Consortium (HRC), UK10K, and 1000 Genomes Phase 3 panels. Genetic principal components and kinship coefficients were centrally estimated by UK Biobank to account for population stratification and relatedness [19, 21].

### Study Variables

The dependent variable in this study was the incidence of stroke. The independent variables included both conventional risk factors and genetic liability to stroke (**Supplementary Table 1**). Independent Variables included the following:

- Sociodemographic factors - age (in years) and sex (male/female).
- Lifestyle factors - alcohol consumption (current, previous, never) and cigarette smoking status (current, previous, never).
- Clinical factors - Body mass index (BMI), total cholesterol (TC) and low-density lipoprotein (LDL) levels, and diabetes mellitus (DM).

DM was defined based on any of the following criteria at baseline: self-reported physician diagnosis of diabetes, (ii) use of insulin or other glucose-lowering medication, (iii) hemoglobin A1c (HbA1c) ≥ 48 mmol/mol (6.5%), or (iv) fasting glucose level ≥ 7.0 mmol/L [22].

### Definition of the Outcome

The primary outcome was the incidence of stroke, as defined by the International Classification of Diseases, 10th Revision (ICD-10 codes I60–I67). Stroke events were identified using Hospital Episode Statistics (HES) records and death registries, capturing first-ever stroke events with the following ICD-10 codes: subarachnoid haemorrhage (I60.0–I60.9), intracerebral haemorrhage (I61.0–I61.9), cerebral infarction (I63.0–I63.9), stroke not specified as haemorrhage or infarction (I64), occlusion and stenosis of pre-cerebral and cerebral arteries (I65.0–I65.9, I66.0–I66.9), and other cerebrovascular diseases (I67.0–I67.9). The follow-up period was calculated from each participant’s baseline health assessment date to the first stroke event or the end of follow-up (March 31, 2017), whichever occurred first. Participants who did not experience a stroke during follow-up were censored at that point.

### Computation of Genetic Liabilities

The selection of single-nucleotide polymorphisms (SNPs) and the preprocessing steps have been described in our previous manuscript [12]. Briefly, SNPs associated with stroke were selected from previously published genome-wide association studies, with the primary source being [23], which focused on stroke genetics in European ancestry populations (Supplementary Data S1). Genetic liability to stroke was quantified using a polygenic risk score (PRS), computed with PLINK 1.9. The PRS was calculated as a weighted sum of stroke-associated risk alleles, where the weights corresponded to the SNP effect sizes (β coefficients) reported in the reference GWAS. Key quality control procedures included:

- Filtering SNPs with low call rates or violations of the Hardy-Weinberg equilibrium,
- Excluding SNPs with minor allele frequency (MAF) < 1%,
- Linkage disequilibrium (LD) pruning using a window size of 250 kb, step size = 50, and an r² threshold of 0.1.

After LD pruning, 252,903 SNPs were retained for PRS calculation (**Supplementary Figure 1**). The PRS was computed using the --score function in PLINK. The resulting PRS and all quantitative independent variables were standardized using the “scale” function in the R package and included as independent variables in regression models to assess the association between genetic predisposition to stroke and incident stroke among individuals with hypertension.

### Statistical Analyses

The statistical analysis approach used in this study follows the methodology described in our previous manuscript [12]. Briefly, baseline characteristics were summarized using descriptive statistics, including frequencies and percentages for categorical variables and means with standard deviations for continuous variables. Differences between stroke and non-stroke groups were assessed using chi-square tests for categorical variables and Wilcoxon rank-sum tests for continuous variables. Baseline characteristics of the study population were summarized using the *gtsummary* and *table1* packages in the R program.

To assess the relationship between the predictors and incident stroke, we used univariable and multivariable Cox proportional hazards regression models (CoxPH). Before multivariable modelling, we checked for multicollinearity among quantitative variables by using Pearson correlation coefficients with the *“cor”* function. Highly correlated variables (r² ≥ 0.80) were identified with the “*findCorrelation”* function from the caret package in R and visualized using the *ggcorrplot* package. Variables with high collinearity and limited univariable association with the outcome were excluded to prevent overfitting (Figure 2).

**Figure 2.**
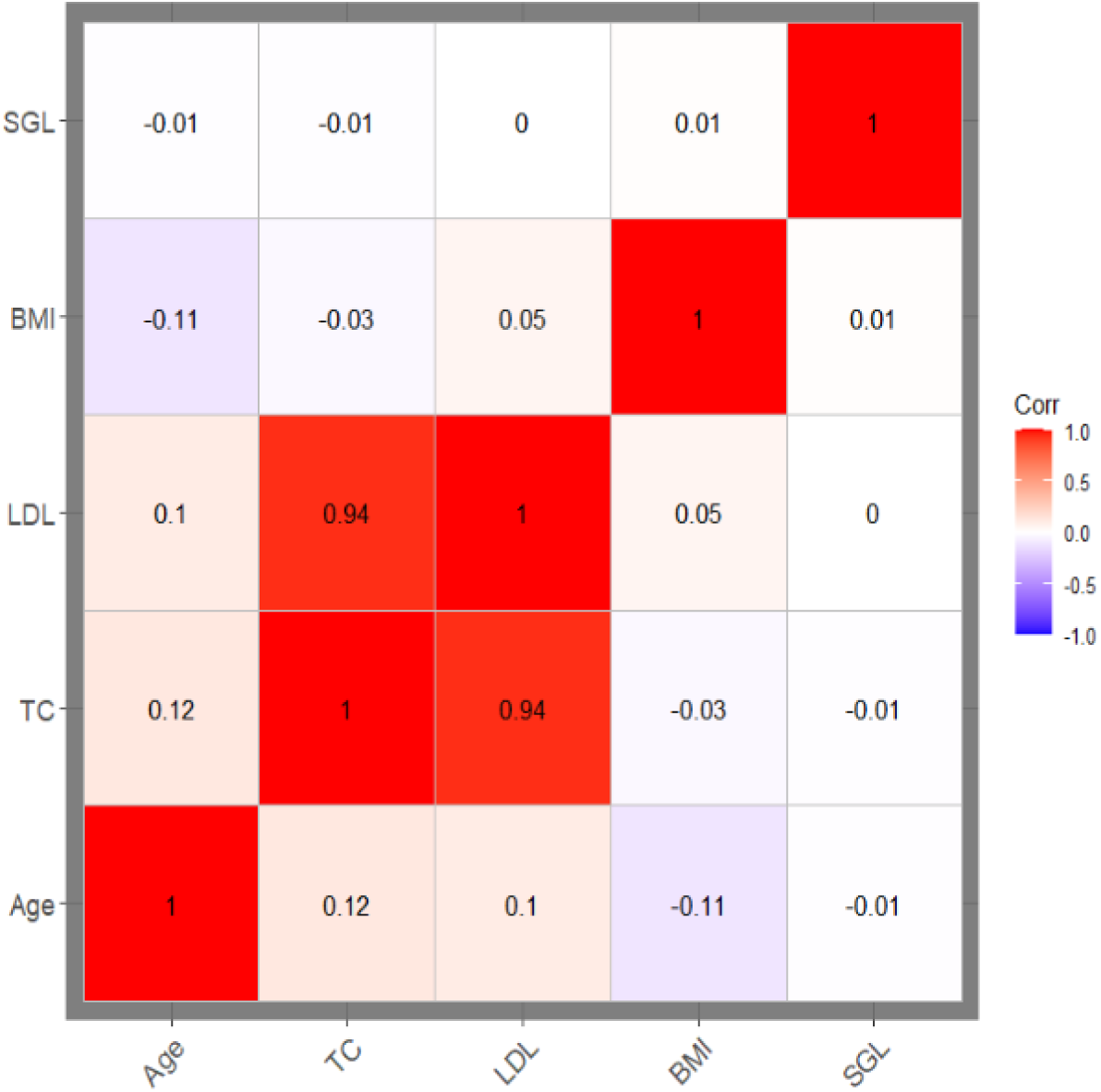
Correlation matrix plot: The plot shows the correlation coefficients between numerical features. TC and LDL are highly correlated (r^2^ >0.8). LDL was excluded from further analysis (prediction model construction). BMI: Body mass index; TC: Total cholesterol; LDL: Low-density lipoprotein cholesterol.

To examine differences in stroke-free survival time among hypertensive participants with varying levels of genetic liability, we performed Kaplan–Meier survival analysis over the defined follow-up period. The survival functions were stratified by categories of genetic liability to stroke (low, intermediate, and high) and compared using the log-rank test (Supplementary Figure 2).

To study the effect of the stroke genetic liability while adjusting for the impact of different covariates, sequences of multivariate CoxPH models were created. Model 1 included only stroke genetic liability, model 2 adjusted for age and sex, model 3 included adjustments for age, sex, diabetes mellitus, and total cholesterol levels, and model 4 included adjustments for age, sex, diabetes mellitus, and total cholesterol, as well as smoking and alcohol status.

The results of the multiple Cox proportional hazards regression analysis are presented as adjusted hazard ratios (HRs) along with the corresponding 95% confidence intervals (CIs). A significance level of p < 0.05 indicates that a predictor is independently associated with the incidence of stroke. A forest plot was employed to visualize the hazard ratios for all covariates using the *“forest_model”* function from the *forestmodel* package.

The Cox proportional hazards assumption was evaluated using the global Schoenfeld residuals test, and individual variable assessments were visualized. Meeting the proportionality assumption indicates the validity of the Cox regression results.

All analyses were performed using R software (version 4.4.1) for Windows (R Development Core Team, 2010).

### Prediction Model Development and Performance Assessment

In this study, two sets of prediction models were developed to predict the incidence of stroke. (1) The conventional risk factors model (model without genetic liability), which combines the conventional risk factors selected from univariable association tests, and (2) the integrated prediction model, which combines genetic liability for stroke (genetic risk) and the conventional risk factors.

Using the “*createDataPartition*” function from the *care*t package, we randomly partitioned our dataset into a training set (70%; N = 81,352; Event = 959 and Non-event = 80,393) and a testing set (30%; N = 34,864; Event = 417 and Non-event = 34,447). The training data was used to create predictive models, while the testing data was utilized to assess the models’ performance.

### Implementation

To assess the added predictive value of stroke genetic liability (SGL) for predicting stroke in hypertensive individuals, we implemented and compared various statistical and machine learning models using UK Biobank data. We implemented both the continuous- and discrete-time survival models to predict the risk of stroke incidence in hypertensive individuals. The continuous-time models included standard Cox proportional hazards (CoxPH) and penalized Cox regression (Coxnet). For penalized logistic regression (GLMNet) and a neural network-based discrete survival model (Nnet), we adopted the discrete-time survival approach. The packages, functions, and parameters used in the implementation of the models are presented in **Supplementary Table 2**. The core concept behind prediction in the discrete-time survival framework is to develop models that estimate the probability of survival within each discrete time interval. By treating the occurrence of an event in each interval as a binary outcome, the task can be framed as a sequence of binary classification problems [24].

In this study, we implemented discrete-time survival analysis by discretizing the continuous follow-up time into defined intervals. This transformation allowed us to restructure the data into a person-period (long) format, in which each row represents a unique individual-time interval combination. This approach ensures that any change in an individual’s status during the follow-up period could be accurately captured. The transformation was conducted using the *“discSurv”* function from the *discSurv* package in R. After reshaping the dataset, we addressed class imbalance, an inherent issue in survival data due to the relatively low incidence of stroke events. To mitigate this, we employed the ROSE (Random Over-Sampling Examples) method using the ROSE function from the ROSE package in R. The use and justification for ROSE were detailed in our previous manuscript [17]. In brief, ROSE generates synthetic examples of the minority class and under-samples the majority class to create a more balanced training set [25]. This pre-processing ensured that the discrete-time models were trained on a balanced dataset, thereby improving model robustness and predictive power, particularly for rare outcomes such as stroke [26].

### Cox Proportional Hazards Model (CoxPH)

Cox proportional hazards regression (Deo, Deo and Sundaram, 2021, Abd ElHafeez *et al.,* 2021) is a popular statistical approach for analysing survival data and determining the association between the time until an event (such as death, failure, or illness recurrence) occurs and one or more predictors. We implemented the Cox proportional hazard models using the *“coxph”* function from the *Survival* package in R software.

In addition to the previously described models, we implemented the following machine learning models in a continuous-time framework: a penalized Cox regression model (CoxNet) and two discrete-time survival models, a penalized logistic regression model, and a neural network, to further explore stroke risk prediction.

### Penalized Regression Models

In comparison to traditional (unpenalized) regression methods, such as Cox and logistic regression, penalized regression models improve prediction performance on new data by applying regularization. Regularization introduces a penalty by shrinking the size of less informative coefficients towards zero and only retaining those with coefficients greater than zero [27, 28]. The penalized method employed in this study was elastic-net, which is a combination of LASSO (L1) and RIDGE (L2) regularization.

#### (1) Penalized Cox Regression (CoxNet)

We utilized an elastic-net regularized Cox regression model with the *“cv. glmnet”* function in the *glmnet* package. The model was trained using 10-fold cross-validation and evaluated by the area under the receiver operating characteristic curve (AUC). The optimized CoxNet model was generated by tuning parameters alpha = 0.5 and lambda = 0.0001 (Supplementary **Table 2).**

#### (2) Penalized Logistic Regression (GLMnet)

We applied an elastic-net regularized logistic regression model using the *“cv. glmnet”* function in the *glmnet* package. The model was trained using 10-fold cross-validation and evaluated by the area under the receiver operating characteristic curve (AUC). The optimized GLMnet model was generated by tuning parameters alpha = 0.5 and lambda = 0.002 **(Supplementary Table 2).**

### Neural Network Model (Nnet)

A feedforward neural network was trained using the Nnet method within the caret framework. The neural net was described and implemented in our previous manuscript [17]. The survival time was discretized into intervals, and the model was evaluated using AUC under 10-fold cross-validation. The optimized neural network model was generated by tuning parameters size = 5 and decay = 0.1 (**Supplementary Table 2).**

The predictions from all the models were obtained using the “*predict”* function. In this study, we additionally focused on probability calibration to improve the interpretability and reliability of stroke risk predictions. To calibrate the predicted probabilities, we applied Platt scaling, also known as the sigmoid method [29] to transform model-generated scores into calibrated probability estimates. This post-processing technique involves fitting a logistic regression model where the model-predicted probabilities are used as the independent variable, and the binary outcome (stroke occurrence) is the dependent variable [30]. The resulting transformation produces rescaled probability estimates that more accurately reflect the true event rates, thereby enhancing clinical interpretability. Details have been described in our previous manuscripts [12, 17].

### Assessing model prediction performance

We assessed the predictive performance of each model in the testing dataset using the receiver operating characteristic curve (AUC) and the Brier Score (BS). We assessed the model’s discrimination ability using the AUC. Model calibration and overall performance were assessed using the BS, which measures the mean squared difference between predicted probabilities and actual outcomes; lower BS values indicate better calibration and prediction accuracy [30]Detailed descriptions of these metrics can be found in our previous work [12, 17].

For the continuous-time survival models, CoxPH and CoxNet, we used the “*rcorr.cens”* function from the *Hmisc* package to calculate the concordance index (c-index or AUC), and for the discrete-time survival models, GLMnet and Nnet, the AUC was calculated using the “*roc”* function from the *pROC* package. Furthermore, we evaluated the discriminative performance of all models specifically in hypertensive individuals at various time points (2, 4, 6, 8, and 10 years) using the “*timeROC”* function from the *timeROC* package. ROC curves were generated for each model using the *“plot”* function.

## RESULTS

### Study Characteristics and Statistical Analysis

**Table 1** presents the baseline characteristics of the study population, composed of 116,216 unrelated participants of European ancestry with hypertension, including 13,766 incident cases and 102,450 controls. Key demographic and clinical features significantly differ between the two groups.

**Table 1:** Baseline Characteristics of Study Population Stratified for Stroke Event and Non-Stroke Event among Hypertension Patients with UK Biobank Participants (N=116,216).

| Characteristics | Overall<br>(N=116,216) | Control<br>(N=102,450) | Case<br>(N=13,766) | HR<br>(95% CI) | p-value |
| --- | --- | --- | --- | --- | --- |
| Age(years);<br>Mean( SD) | 57.6<br>(7.53) | 57.6<br>(7.53) | 60.9<br>(6.70) | 1.67<br>(1.57, 1.78) | <0.001 |
| Body mass index<br>(kg/m<sup>2</sup>);<br>Mean (SD) | 28.0<br>(4.83) | 28.0<br>(4.83) | 27.9<br>(4.77) | 0.98<br>(0.93, 1.03) | 0.44* |
| Total cholesterol<br>(mmol/L);<br>Mean (SD) | 6.05<br>(1.06) | 6.05<br>(1.06) | 5.96<br>(1.07) | 0.92<br>(0.88, 0.97) | 0.004 |
| Low-density lipoprotein<br>(mmol/L);<br>Mean (SD) | 3.84<br>(0.81) | 3.84<br>(0.81) | 3.81<br>(0.81) | 0.96<br>(0.91, 1.02) | 0.16* |
| Sex (Male);<br>n (%) | 55269<br>(47.6%) | 54476 (47.4%) | 793 (57.6%) | 1.50<br>(1.35, 1.67) | <0.001 |
| Diabetes Mellitus (Yes);<br>n (%) | 4638<br>(4.0%) | 4545<br>(4.0%) | 93<br>(6.8%) | 1.75<br>(1.42, 2.16) | <0.001 |
| **Smoking status** |  |  |  |  | <0.001 |
| Current n (%) | 37098<br>(31.9%) | 36579 (31.9%) | 519<br>(37.7%) | 1.33<br>(1.19, 1.48) |  |
| Previous; n (%) | 1320<br>(1.1%) | 1283<br>(1.1%) | 37<br>(2.7%) | 1.15<br>(0.83, 1.59) |  |
| **Drinking status** |  |  |  |  | <0.001 |
| Current; n (%) | 108867<br>(93.7%) | 107635<br>(93.7%) | 1232 (89.5%) | 0.61<br>(0.49, 0.78) |  |
| Previous; n (%) | 3333<br>(2.9%) | 3263<br>(2.8%) | 70<br>(5.1%) | 2.67<br>(1.92, 3.72) |  |
| n (%); Mean (SD), HR = Hazard Ratio, CI = Confidence Interval |  |  |  |  |  |

The p-value is from a univariate analysis of the Cox Proportional Hazard Model comparing the distribution of the baseline characteristics among stroke events and non-stroke events in hypertension patients. \*Not significant; **DM** = Diabetes Mellitus, **HR** = Hazard Ratio, CI = Confidence Interval, **SD** = Standard Deviation.

Participants who developed a stroke during the study were, on average, older than controls (mean age 60.9 vs. 57.6 years). Age was strongly associated with incident stroke (HR = 1.67, 95% CI = [1.57, 1.78], *p-value* < 0.001). Male participants had a significantly higher hazard than female participants (HR = 1.50, 95% CI = [1.35, 1.67], *p-value* < 0.001). Among lifestyle factors, current smoking was associated with a higher risk of stroke (HR = 1.33, 95% CI = [1.19, 1.48], *p-value* < 0.001), while current alcohol consumption was inversely associated with the risk of stroke (HR = 0.61, 95% CI = [0.49, 0.78], *p-value* < 0.001).

Among the clinical factors, diabetes mellitus was also strongly associated with an increased risk of incident stroke (HR = 1.75, 95% CI = [1.42, 2.16], *p-value* < 0.001). Higher total cholesterol was marginally protective (HR = 0.92, 95% CI: 0.88–0.97, p = 0.004), whereas low-density lipoprotein (LDL) cholesterol and BMI were not significantly associated with event risk (*p-value* = 0.16 and 0.44, respectively).

The correlation matrix plot showed that total cholesterol (TC) and low-density lipoprotein (LDL) are highly correlated, and the stroke genetic liability is independent of the other covariates (**Figure 2**)

**Table 2** shows the results of a sequence of Cox proportional hazards models evaluating the relationship between stroke genetic liability (as a continuous variable) and incident stroke, from univariable (Model 1) to adjusted multivariable models (Models 2–4). Genetic liability was consistently associated with a statistically significant increased risk of stroke.

**Table 2:**
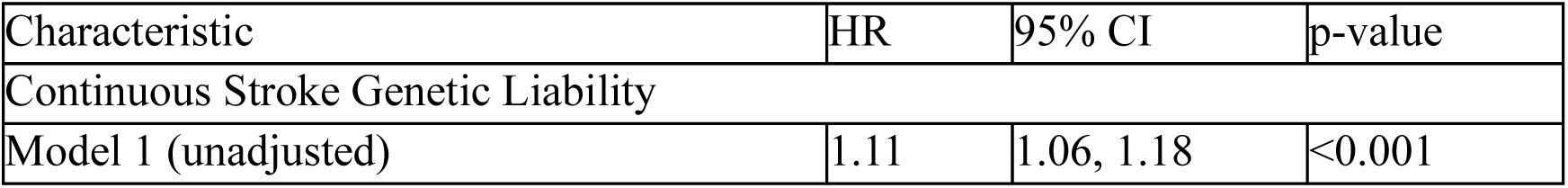
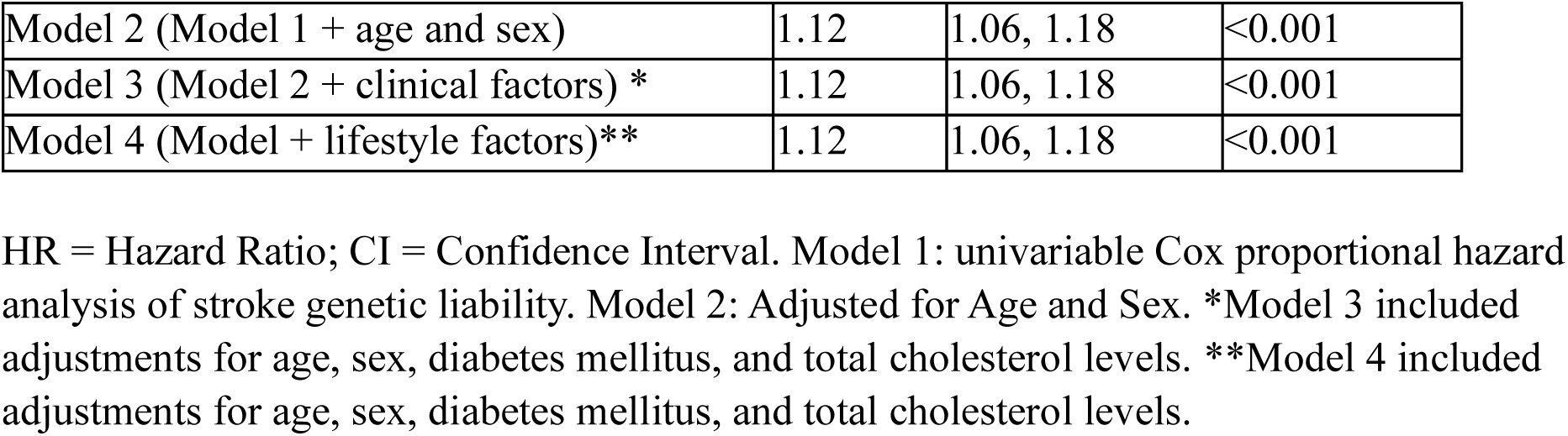
Univariable and Multivariable Cox Proportional Hazard Analysis Assessing the Association of Stroke Genetic Liability with Stroke.

In the unadjusted model (Model 1), the hazard ratio (HR) was 1.11 (95% CI = [1.06, 1.18], *p-value* < 0.001), indicating that for each unit (standard deviation) increase in genetic liability, the risk of stroke increased by approximately 11%. This association remained steady after adjustment for age and sex in Model 2 (HR = 1.12, 95% CI = [1.06, 1.18], *p-value* < 0.001) and was unchanged with further inclusion of conventional risk factors such as diabetes, body mass index, and total cholesterol in Model 3. Even after including additional covariates such as smoking status and alcohol status in Model 4, the effect estimate remained stable (HR = 1.12, 95% CI = [1.06, 1.18], *p-value* < 0.001).

These results suggest that stroke genetic liability is an independent risk factor for stroke risk, showing a consistent and statistically significant effect across all levels of adjustment in the models. The assessment of the Cox proportional hazards assumption indicated that all the predictor variables individually achieved a *p-value* **>** 0.05 based on Schoenfeld residuals, and the global test also established that the overall model satisfied the assumption (*p-value* **>** 0.05). These results suggest that the Cox proportional hazards assumption was not violated for any predictor variable or the model as a whole (Supplementary **Table 3 and Supplementary Figure 3**).

### Prediction Performance of the Models

**Table 3** presents the results of performance from a series of prediction model techniques employed in this study

**Table 3:** The Results of the Model Performance for Incident Stroke Prediction in the Hypertensive Patients in UK Biobank.

| Models | Model Features | Sample | AUC (95% CI) | Brier Score |
| --- | --- | --- | --- | --- |
| CoxPH |  |  |  |  |
|  | Conventional factors + genetics | whole population (n =116,216) | 67.0 (64, 70) | 0.01 |
|  | Conventional factors + genetics | Men (n = 55,269) | 68.0 (64, 71) | 0.01 |
|  | Conventional factors + genetics | Women (n = 60,947) | 65.0 (60, 69) | 0.01 |
|  | Conventional factors + genetics | Age > 59 (n = 55370) | 60.0 (56, 64) | 0.02 |
|  | Conventional factors + genetics | Age < 59 (n = 60846) | 58.0 (53, 63) | 0.02 |
| **CoxNet** |  |  |  |  |
|  | Conventional factors + genetics | whole population (n=116,216) | 67.0 (64, 70) | 0.01 |
|  | Conventional factors + genetics | Men (n = 55,269) | 67.0 (64, 71) | 0.01 |
|  | Conventional factors + genetics | Women (n = 60,947) | 65.0 (60, 69) | 0.01 |
|  | Conventional factors + genetics | Age > 59 (n = 55370) | 60.0 (57, 64) | 0.02 |
|  | Conventional factors + genetics | Age < 59 (n = 60846) | 56.0 (51, 61) | 0.01 |

**Table 3 Continued**
| **Models** | **Model Features** | **Sample** | **AUC (95% CI)** | **Brier Score** |
| --- | --- | --- | --- | --- |
| **GLMnet** |  |  |  |  |
|  | Conventional factors + genetics | whole population (n=116,216) | 67.0 (64, 70) | 0.001 |
|  | Conventional factors + genetics | Men<br>(n = 55,269) | 67.0<br>(63, 70) | 0.002 |
|  | Conventional factors + genetics | Women<br>(n = 60,947) | 65.0<br>(61, 69) | 0.001 |
|  | Conventional factors + genetics | Age > 59<br>(n = 55370) | 61.0<br>(58, 65) | 0.002 |
|  | Conventional factors + genetics | Age < 59<br>(n = 60846) | 57.0<br>(52, 62) | 0.001 |
| **Nnet** |  |  |  |  |
|  | Conventional factors + genetics | whole population<br>(n=116,216) | 66.0<br>(63, 69) | 0.001 |
|  | Conventional factors + genetics | Men<br>(n = 55,269) | 65.0<br>(62, 69) | 0.002 |
|  | Conventional factors + genetics | Women<br>(n = 60,947) | 64.0<br>(59, 68) | 0.001 |
|  | Conventional factors + genetics | Age > 59<br>(n = 55370) | 60.0<br>(56, 63) | 0.002 |
|  | Conventional factors + genetics | Age < 59<br>(n = 60846) | 55.0<br>(50, 60) | 0.001 |

Across the full study sample (N = 116,216), the inclusion of genetic liability of stroke yielded small but statistically significant improvements in the risk of stroke prediction.

Both the Cox proportional hazards (CoxPH) and penalized Cox regression (CoxNet) achieved an equal AUC of 67.0 (95% CI = [64, 70]). The Brier score (BS) for both models was 0.01. The penalized logistic model (GLMnet) also achieved an AUC of 67.0 (95% CI = [64, 70]), while the neural network-based model (Nnet) achieved an AUC of 66.0 (95% CI = [63, 69]). Both GLMnet and Nnet achieved a BS of 0.001. The AUC and BS values indicate good prediction performance across all the models.

Our analysis of hypertensive men and women separately shows that the model performed better in hypertensive men than in hypertensive women.

Among hypertensive men (n = 55,269), the CoxPH achieved an AUC of 68.0(95% CI = [64, 71]) and BS of 0.01. The model achieved an AUC of 67.0 (95% CI = [64, 71]) and BS of 0.01 for CoxNet, an AUC of 67.0 (95% CI = [63, 70]) and BS of 0.002 for GLMnet. The Nnet attained an AUC of 65.0(95% CI = [62, 69]) BS of 0.002. Among hypertensive women participants (n = 60,947), both the CoxPH and CoxNet achieved an equal AUC of 65.0 (95% CI = [60, 69]) and equal BS of 0.01. GLMnet achieved an AUC of 65.0 (95% CI = [61, 69]) and BS of 0.001, while Nnet achieved an AUC of 64.0 (95% CI = [59, 68]) and BS of 0.001

The models had a better discrimination performance for participants who were older than the average age (59 years old) of the whole study sample than for younger participants. In older participants (n=55,370), the GLMnet achieved an AUC of 61.0 (95% CI = [58, 65]) and BS of 0.002, while CoxPH, CoxNet, and Nnet achieved a similar AUC of 60.0.

### Model Performance at Multiple Follow-up Times Points

**Table 4** presents the discriminative abilities of the models at different follow-up time points (2, 4, 6, 8, and 10 years) for the whole study sample. The result shows that all the continuous-time survival models’ performances improved with time. CoxPH and CoxNet performed similarly and consistently (approx.65 to 67 AUC) across the follow-up time points. CoxNet slightly improved with time and performed best in year 8 (AUC = 67.37). The discrete-time survival models (GLMnet and Nnet) performed strongly in years 4 and 6, then degraded after. Both GLMnet and Nnet achieved their highest AUCs in year 4 (AUC = 69.98 and 68.78, respectively), suggesting strong mid-term prediction.

**Table 4:** Model Performance at Multiple Follow-Up Time Points in the Testing Set. Table 4 shows the time-dependent ROC AUC (Area Under the Curve) estimates over different years (2, 4, 6, 8, 10) using IPCW (Inverse Probability of Censoring Weighting) for six predictive models: CoxPH, CoxNet, GLMnet, and Nnet.

| Years | Cases | Survivors<br>(without<br>the event) | Censored | CoxPH<br>(AUC %) | Coxnet<br>(AUC %) | GLMnet<br>(AUC %) | Nnet<br>(AUC %) |
| --- | --- | --- | --- | --- | --- | --- | --- |
| 2 | 75 | 34789 | 0 | 64.62 | 64.63 | 63.18 | 62.47 |
| 4 | 168 | 34696 | 0 | 65.26 | 65.29 | 69.98 | 68.78 |
| 6 | 282 | 34582 | 0 | 65.91 | 65.91 | 68.68 | 67.10 |
| 8 | 389 | 20450 | 14025 | 67.35 | 67.37 | 66.72 | 65.21 |
| 10 | 417 | 0 | 34447 | N/A | N/A | 50.72 | 53.40 |

## DISCUSSIONS

This large-scale cohort study investigated the predictive value of genome-wide stroke genetic liability derived from 252,903 stroke-associated SNPs in over 116,000 hypertensive individuals of European ancestry. In the current study, we constructed machine learning models and assessed their ability to predict stroke incidents in all hypertensive participants and stratified by age and sex using a survival analysis framework. Our models included traditional risk factors of stroke alongside genetic liability. To examine the discriminatory accuracy of time-to-event data models, we employed both discrete- and continuous-time machine learning algorithms. These included Elastic Net regression models, specifically the penalized Cox model (CoxNet) and penalized logistic regression (GLMnet), as well as a neural network (Nnet). These models were compared to the Cox proportional hazards (CoxPH) model, which is used as the gold standard for survival analysis.

Our **main findings** were

(1) In all hypertension patients, all the models (both discrete and continuous-time survival models) showed modest discriminating value, with AUC ranging from 66 to 68. CoxNet and GLMnet achieved equal discrimination ability as the CoxPH model in the follow-up period. The discrete-time survival models (GLMnet and Nnet) outperformed the continuous-time models at narrower follow-up intervals (2-4 years and 4-6 years from baseline). (2) All models consistently achieved higher AUC values in hypertensive men than in hypertensive women. (3) All models consistently achieved higher AUC values in older participants (over the median age of 59 years old) than in younger participants.

In comparison with our previous studies on the stroke risk prediction [12], the current study (1) focuses on hypertensive individuals. (2) While our previous work focused on models using continuous-time survival models, the current work adds discrete-time machine learning models (GLMnet, a form of penalized logistic regression model, and Nnet). (3) Discretized the follow-up time and incorporated it as a covariate within the discrete time machine learning models (4) Our present work additionally extended and leveraged the readily available open-source software packages in R-program for the binary classification problem to survival analysis frameworks. That is, we reformulated the continuous-time survival model as a binary classification problem. This approach overcomes several limitations of traditional continuous-time survival models, including problems with tied event times, the proportional hazard assumption requirement. Discrete-time survival models offer practical alternatives to continuous-time models, particularly when the exact event times are unknown, and only the time interval during which the event occurs is known. This framework restructures survival analysis as a series of binary classification problems, enabling a wide range of classification algorithms to be used for estimating conditional survival probabilities [31].

While machine learning methods are appropriate for capturing complex, non-linear relationships between predictors and outcomes, our findings suggest that the Elastic Net models (GLMnet and CoxNet) performed similarly to our reference regression model (CoxPH) for stroke prediction in hypertensive patients. In contrast, the more complex model, Neural Network (Nnet), demonstrated inferior performance relative to CoxPH. This observation suggests that the CoxPH model may be sufficient for developing an effective and accurate stroke risk prediction model in hypertensive individuals from the UK Biobank (UKB). This observation could be due to the absence of non-linear or complex interactions among the predictor variables used in the current study.

This finding is consistent with the results of our previous work [12] when we demonstrated that CoxPH outperformed all the machine learning models, including Random Forest (RF), the Gradient Boosting model (GBM), and the Decision Tree (DT). However, in our previous work, we did not consider the binary classification machine learning method and did not explore age and sex strata.

### Comparison between traditional and machine learning models that include the effect of time

The performance of Machine learning techniques has been compared to traditional Cox regression models in predicting stroke risk; however, the results in the literatures are inconsistent. For example, two previous studies, Chen, Y [11] and Chun [9] each created ML-based prediction models and compared their performance against Cox regression. While both studies highlighted the potential of ML techniques, their conclusions about superiority to Cox regression were not conclusive.

Chun and colleagues [9] with age, hypertension, coronary heart disease, diabetes, and smoking and observed that for the 9-year risk of stroke prediction in Chinese adults, the Cox regression model and the machine learning models had similar prediction performance. The Cox regression model marginally outperformed the Random Survival Forest model but performed worse than the Gradient Boosted Trees.

For predicting 30-day stroke readmission in a Taiwanese cohort, Chen, Y, and colleagues included a wide range of clinical, demographic, and pre-rehabilitation functional status scores in their prediction models. They observed that among the machine learning models, only the neural network and the random forest models outperformed the Cox regression model. These results differed from the findings of our current study, which found that both the neural network and random forest models underperformed in comparison to the CoxPH model.o the CoxPH model.

Unlike our study, the two previous studies were conducted in Asian populations, whose genetic architectures differ from the European population we studied. While [11] explored the prediction of 30-day stroke readmission and Chun [9] focused on the prediction of first-ever incident stroke within a population-based cohort, our study focussed on predicting first-time stroke events in hypertensive Europeans beyond the 30-day window and uniquely incorporated stroke genetic liability as a predictor. The methodological divergences highlight the uniqueness of our approach and may explain differences in prediction accuracy and interpretability between studies. These fundamental differences in research methodology, outcome definitions, and study population restrict direct comparisons of our findings with the previous studies and may explain the difference in model performances.

### Comparison based on the different follow-up time lengths

When we evaluated model performance across different follow-up intervals for hypertensive individuals, we observed that during shorter follow-up intervals (2–4 years and 4–6 years from baseline), all the discrete-time survival models outperformed the continuous-time survival models. Within shorter follow-up intervals, the GLMnet was the best-performing model, followed by the Nnet model. In contrast, with a longer follow-up interval (6–8 years), the continuous-time survival models performed better, with Cox proportional hazards (CoxPH) and CoxNet emerging as the best-performing models. These observations were consistent with the findings of Chun [9] who demonstrated that the binary classification machine learning models, including logistic regression (LR), support vector machines (SVM), gradient boosting trees (GBT), and multilayer perceptron (MLP), outperformed survival models such as Cox regression and random survival forests. They reported the best performances in predicting stroke were observed within shorter follow-up intervals (0–3 years, 3–6 years, and 6–9 years from baseline). This implies that for prediction within shorter follow-up times, discrete-time survival models (GLMnet and Nnet) are more suitable for risk prediction.

### Gender differences

Across all models, both machine learning and traditional models, the model’s discrimination ability was consistently better among hypertensive men compared to hypertensive women. Among hypertensive men, the CoxPH model provided better discriminative accuracy than machine learning models. These findings diverged from the previous study by of Chun [9].

Within a population-based Chinese cohort, Chun [9] compared the prediction performance of machine learning algorithms with the Cox regression model for predicting the first-ever stroke in men and women separately. They observed that the models improved stroke prediction in women more than in men, with GBT providing the best discrimination (AUC: 0.833 in men, 0.836 in women) and calibration. This observation deviates from the findings of our current study, in which both machine learning and Cox model improved stroke prediction in hypertensive men more than in hypertensive women, with the CoxPH providing the best discrimination (AUC: 68 % in men, 65% in women). This further highlights the possible impact of population characteristics, outcomes, and study methodology on predictive model performance.

### Age differences

Across all models, both machine learning and traditional models, the discrimination performance of the models consistently achieved a better discrimination value among older hypertensive participants compared to younger hypertensive participants. Among older hypertensive participants, the GLMnet model provided better discriminative accuracy. Our finding was in consistent with findings from several studies, including Ding [32] and Gong [14], which demonstrated that the risk of stroke increases with age, especially among older hypertensive patients. These observations show the potential age-based differences in model performance and/or underlying risk factor patterns.

The general clinical implication of our study is that the absolute risk estimates derived from the stroke genetic liability-enhanced model could help identify older hypertensive patients, especially hypertensive men, at high risk of stroke, who can then be prioritized for preventive interventions, including the start of pharmacological treatments. A study of 100,000 UK adults [33] found that the small improvement in C-index by polygenic risk scores for cardiovascular disease (CVD) could translate to a 7% increase in CVD event prevention compared to conventional risk factors alone.

### Strengths and Limitations

Our study has several strengths. First, this is the first study to employ both discrete and continuous-time survival models to evaluate the added predictive value of stroke genetic liability and predict risk of stroke in hypertensive individuals of European ancestry. Second, our study utilized a large sample size and a long follow-up time to accurately identify stroke incident cases. However, there are some limitations in our study. First, we incorporated few predictor variables into our models. Second, data on lifestyle factors such as drinking and smoking behaviours are self-reported and may be inaccurate. Third, the small number of incident stroke cases in our study may have an impact on the predictive power of the models. Therefore, we recommend that future study should include a large number of stroke occurrence cases and a broader set of predictor variables to improve the efficacy of the machine learning models over the CoxPH model.

## Conclusion

In conclusion, within the survival analysis framework, the Cox proportional hazards (CoxPH) model demonstrated superior performance in predicting stroke risk among hypertensive patients compared to the machine learning models evaluated. Our findings may provide a stroke risk stratification and prevention of stroke in the hypertensive population.

## Data Availability

All data produced in the present study are available upon reasonable request to the authors

## Acknowledgment

This research has been conducted using the UK Biobank Resource under Application Number 60549. The MEGASTROKE project received funding from sources specified at http://www.megastroke.org/acknowledgments.html”.

## Author contributions

Conceptualization, R.P.; Data curation, Formal analysis, G.M.; Investigation, G.M.; Methodology, G.M., R.P.; Project administration, G.M., and R.P.; Resources, R.P.; Supervision, R.P; Writing— original draft, G.M.; Writing—review & editing, G.M., and R.P. All authors have read and agreed to the published version of the manuscript.

## Institutional Review Board Statement

The study was conducted in accordance with the Declaration of Helsinki and approved by the Institutional Review Board (or Ethics Committee) of Brunel University London, College of Health, Medicine, and Life Sciences (27675-LR-Feb/2021-31174-2, 5 February 2021)

## Informed Consent Statement

Informed consent was obtained from all subjects involved in the study.

## Data Availability Statement

Not applicable.

## Supplementary Materials

**Supplementary Data S1-** Supplementary Data S1: List of genetic variants summary statistics used to construct the genetic risk scores.

